# Reconnecting the Hand and Arm to the Brain: Efficacy of Neural Interfaces for Sensorimotor Restoration after Tetraplegia

**DOI:** 10.1101/2023.04.24.23288977

**Authors:** Eric Z. Herring, Emily L. Graczyk, William D. Memberg, Robert D. Adams, Guadalupe Fernandez Baca-Vaca, Brianna C. Hutchison, John T. Krall, Benjamin J. Alexander, Emily C. Conlan, Kenya E. Alfaro, Preethi R. Bhat, Aaron B. Ketting-Olivier, Chase A. Haddix, Dawn M. Taylor, Dustin J. Tyler, Robert F. Kirsch, A. Bolu Ajiboye, Jonathan P. Miller

## Abstract

**Background:** Paralysis after spinal cord injury involves damage to pathways that connect neurons in the brain to peripheral nerves in the limbs. Re-establishing this communication using neural interfaces has the potential to bridge the gap and restore upper extremity function to people with high tetraplegia.

**Objective:** We report a novel approach for restoring upper extremity function using selective peripheral nerve stimulation controlled by intracortical microelectrode recordings from sensorimotor networks, along with restoration of tactile sensation of the hand using intracortical microstimulation.

**Methods:** A right-handed man with motor-complete C3-C4 tetraplegia was enrolled into the clinical trial. Six 64-channel intracortical microelectrode arrays were implanted into left hemisphere regions involved in upper extremity function, including primary motor and sensory cortices, inferior frontal gyrus, and anterior intraparietal area. Nine 16-channel extraneural peripheral nerve electrodes were implanted to allow targeted stimulation of right median, ulnar (2), radial, axillary, musculocutaneous, suprascapular, lateral pectoral, and long thoracic nerves, to produce selective muscle contractions on demand. Proof-of-concept studies were performed to demonstrate feasibility of a bidirectional brain-machine interface to restore function of the participant’s own arm and hand.

**Results:** Multi-unit neural activity that correlated with intended motor action was successfully recorded from intracortical arrays. Microstimulation of electrodes in somatosensory cortex produced repeatable sensory percepts of individual fingers for restoration of touch sensation. Selective electrical activation of peripheral nerves produced antigravity muscle contractions. The system was well tolerated with no operative complications.

**Conclusion:** The combination of implanted cortical electrodes and nerve cuff electrodes has the potential to allow restoration of motor and sensory functions of the arm and hand after neurological injury.

## INTRODUCTION

SCI affects over 250,000 people nationwide with over 12,000 new cases each year and less than a 1% full recovery rate^1^. In particular, 52% of all SCI occurs at high cervical levels, resulting in tetraplegia – loss of functional use of all four limbs – and leading to lower quality of life from the inability to independently perform standard activities-of-daily-living (ADL), such as grasping objects for drinking and self-feeding. Many assistive technologies have been developed that provide these individuals with improved abilities to interact with their environment^2–4^. However, control of these devices is limited by inadequate command signals for telling the device what to do, and hence restoration of dexterous function has been elusive. In the case of upper motor neuron injuries, nerves and muscles distal to the injury, with intact lower motor neurons, are physiologically normal and remain electrically excitable^5^, which opens the possibility of using electrical stimulation to reanimate the limb and restore function. Functional electrical stimulation (FES) involving direct stimulation of peripheral nerves and muscles allows for reanimation of paralyzed limbs. This approach has emerged as a powerful technique to allow restoration of function in people with chronic weakness or paralysis^6–10^.

Prior studies into brain-machine interfaces (BMI) have successfully recorded neural activity from motor regions of the brain, and extracted features that can be decoded into multidimensional command signals^11–16^. The combination of BMIs (for a command signal) with FES (to allow movement of the individual’s paralyzed limb) is an appealing solution since it bypasses the injury and restores function using the most intuitive control source and output system conceivable: the individual’s own brain and limbs^16,17^. While these prior BMI-controlled FES studies demonstrated basic feasibility, the achievable functionality was limited in part by the lack of muscle selectivity and/or the limited strength of contractions generated by the FES electrode technology used^16,17^. In addition, prior systems did not attempt to provide sensory feedback about object interactions during BMI-controlled FES grasping. Touch feedback is critical for dexterous object manipulation, and will likely further enhance upper extremity function provided by the BMI-FES system, as has been observed with robotic upper extremity systems^18–20^. In this report, we describe the surgical approach to implantation of percutaneous multi-array intracortical BMI and peripheral nerve FES systems in the same individual with the goal of restoring brain-controlled upper extremity movements. We also present data demonstrating feasibility of each system component. When integrated into a bidirectional sensorimotor system, the ReHAB technology may provide some degree of functional independence to people living with paralysis of the upper extremities.

## METHODS

### ReHAB System Overview

The “Reconnecting the Hand and Arm to the Brain” (ReHAB) system creates a bidirectional computerized interface between sensorimotor regions of the brain and the peripheral nerves, circumventing the spinal injury to create an alternative pathway for transmitting activity between the brain and periphery. A motor pathway records neural activity from electrodes distributed across motor regions of the brain, decodes this information to determine intended movements, and then stimulates sub-fascicular regions of peripheral nerves to actuate appropriate muscles. In parallel, a sensory pathway provides electrical stimulation to the somatosensory cortex, resulting in restored touch percepts. A system overview is shown in Figure 1.

**Figure 1.**
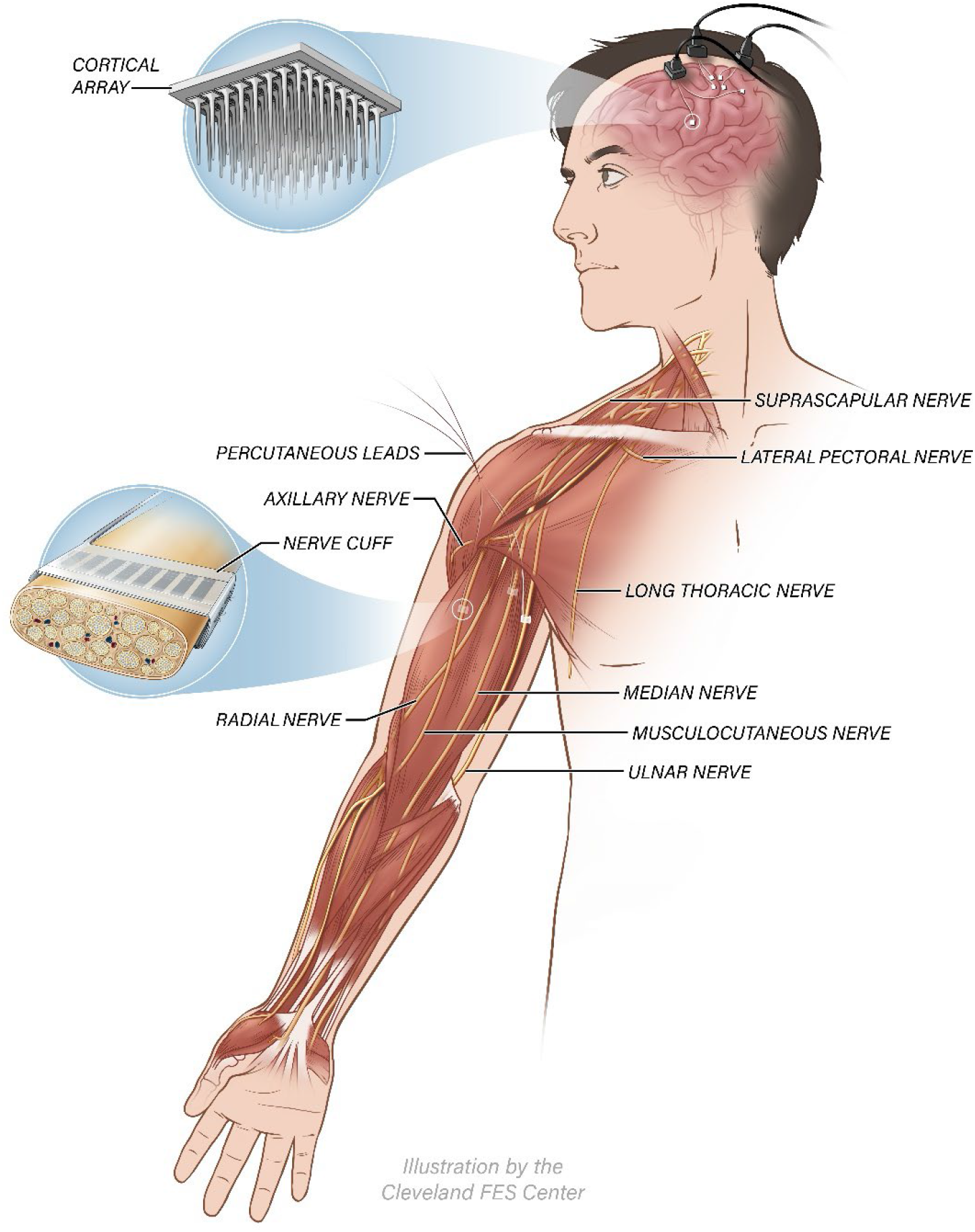
Schematic of the ReHAB system. Cortical arrays implanted in motor regions record brain activity associated with intended movements, which is translated into stimulation patterns sent to nerve cuff electrodes surrounding nerves in the contralateral upper limb. Connections between external neurostimulation hardware and the implanted nerve cuff electrodes are made via percutaneous leads exiting the body in the deltoid and cervical regions. Cortical arrays implanted in somatosensory cortex can record neural activity associated with touch stimuli applied to the hand, and/or elicit sensations of touch on the hand and arm due to stimulation applied through individual electrode contacts.

### Study Participant

A right-handed man in his 20s with C3-C4 AIS B tetraplegia from a diving accident five years prior was enrolled into the study. He had initially undergone a prolonged hospital course involving cervical decompression and stabilization surgery as well as a tracheostomy, but at the time of study enrollment was independent from the ventilator without tracheostomy. An Investigational Device Exemption (ClinicalTrials.gov ID: NCT03898804) was obtained from the FDA for use of intracortical and peripheral nerve electrodes for research purposes. The protocol was reviewed and approved by the University Hospitals Cleveland Medical Center Institutional Review Board. Informed consent was obtained for study enrollment and prior to all procedures.

### Cranial Electrode Implantation

Six 8×8 (64-channel) iridium oxide microelectrode arrays (penetrating 1.5 mm into cortical tissue) were implanted into motor and sensory regions of the left (language dominant) hemisphere. Under awake anesthesia, a bicoronal incision with mobilization of the left temporalis muscle was used to maximize cranial exposure and minimize sacrificing surrounding blood supply. After a left frontoparietal craniotomy, frameless stereotaxis (StealthStation S8 Surgical Navigation System Medtronic PLC, Dublin, Ireland) was used to identify structures of interest, and the central sulcus at the face was confirmed using phase reversal. The hand area on the precentral gyrus was identified through anatomical landmarks^21,22^. The hand-related area of primary somatosensory cortex was then identified through awake sensory mapping, in which four of the five fingers were localized (Figure 2A-B). This information was used to allow placement of two arrays each in primary motor and sensory hand areas. Additional electrodes were implanted in anterior intraparietal area according to landmarks previously described^23^ and in inferior frontal gyrus in areas confirmed to be away from language function using awake speech mapping (Figure 2A-B). Each array was implanted using a high-speed pneumatic press and implantation of grounding wires as previously described^16,24^. The arrays were attached in pairs to three percutaneous pedestals (NeuroPort System, Blackrock Microsystems, Salt Lake City, UT) - one anterior and two posterior to the craniotomy. Connectors were spaced appropriately to allow attachment of headstages for intracortical recording and/or stimulation (Figure 2C-D).

**Figure 2.**
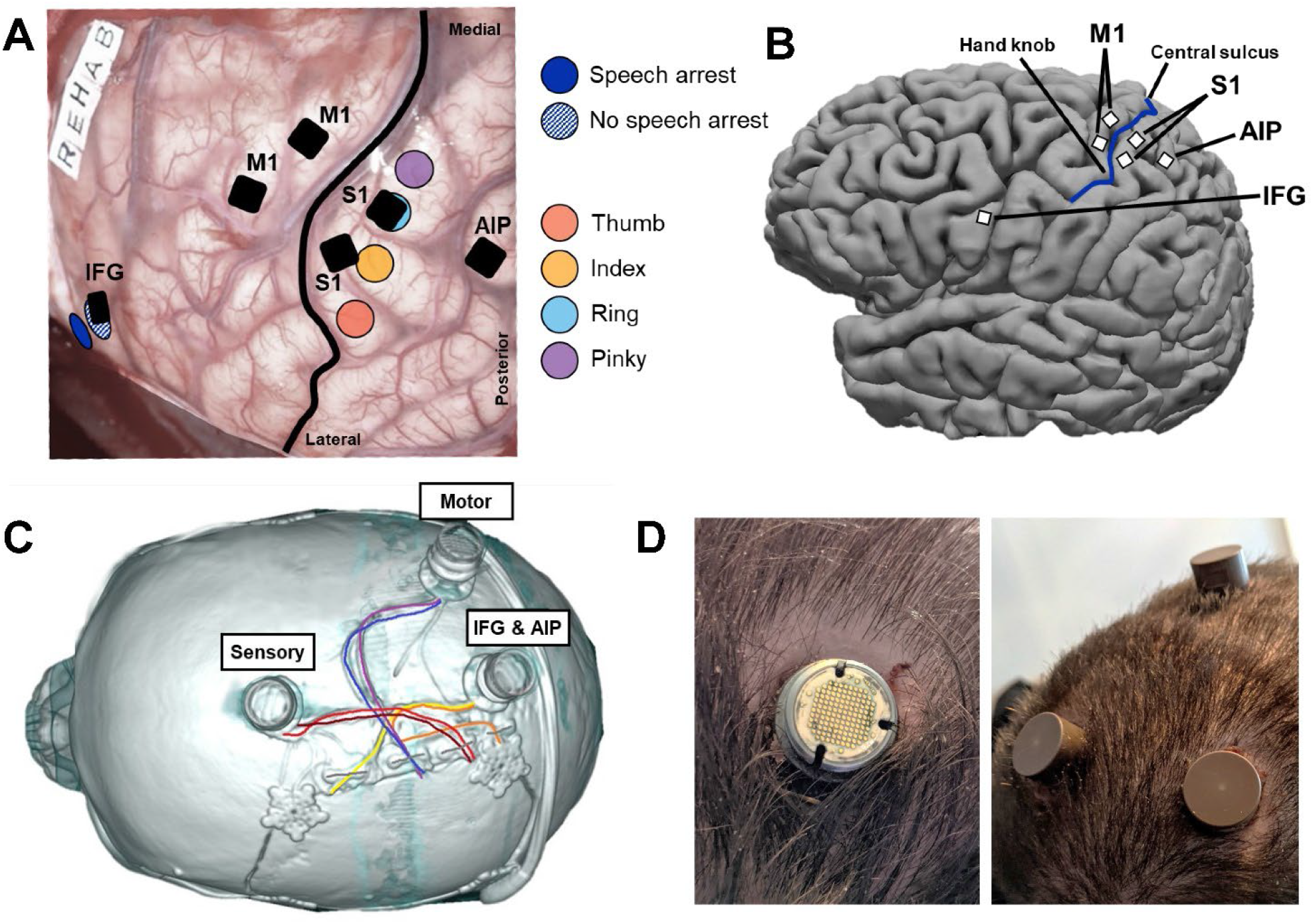
Surgical implantation of the cortical electrodes. (A) Awake stimulation mapping was performed to identify implant locations. Non-speech regions of inferior frontal gyrus (IFG) (striped dark blue) were identified posterior to regions that resulted in speech arrest during stimulation (dark blue). Hand regions of primary somatosensory cortex were located by reports of perceived sensations on individual fingers (red, orange, light blue, purple) during stimulation. Locations selected for the implanted arrays are indicated as black squares. (B) Locations of implanted electrodes on the brain, superimposed upon pre-operative structural MRI. S1 arrays targeted index and ring fingertip locations from (A). M1 arrays were placed directly across the central sulcus, targeting hand and arm area. The IFG array targeted the border of Area 44 and ventral premotor (PMv) cortices, and the AIP array targeted the medial junction of parietal and postcentral sulci. C) CT image of array pedestal locations with associated cabling to implanted arrays. (D) Post-op image of healed array pedestal exit sites with and without caps (in place when system not in use).

### Peripheral Nerve Electrode Implantation

In separate operations under general anesthesia, nine 16-channel composite flat interface nerve electrodes (C-FINEs) were implanted around nerves responsible for dexterous function of the right (dominant) upper extremity. The C-FINE is an extraneural electrode designed to reshape nerves in order to more selectively recruit interior fascicles^25^. Standard surgical approaches were used to provide exposure to the infraclavicular brachial plexus (for median, musculocutaneous, lateral pectoral, proximal ulnar, and long thoracic nerves), distal upper arm (distal ulnar nerve), and posterior upper arm/shoulder (radial, axillary, and suprascapular nerves). High frequency ultrasound was used to identify and evaluate the exposed nerve segments, including size of the nerves, branch points, and number of fascicles. For each nerve, a 16-channel C-FINE was wrapped around the nerve and sutured closed^26^ (Figure 3A), and individual contacts were stimulated intraoperatively to confirm muscle contractions. Percutaneous inline connectors (Figure 3B) were connected and tunneled to percutaneous exit sites located at the lateral deltoid and superclavicular regions (Figure 3C). Some of the percutaneous leads were truncated prior to externalization such that a total of 138 double-wound percutaneous leads passed through 71 exit sites, inclusive of a triceps intramuscular electrode inserted during a subsequent surgery to further enhance arm extension.

**Figure 3.**
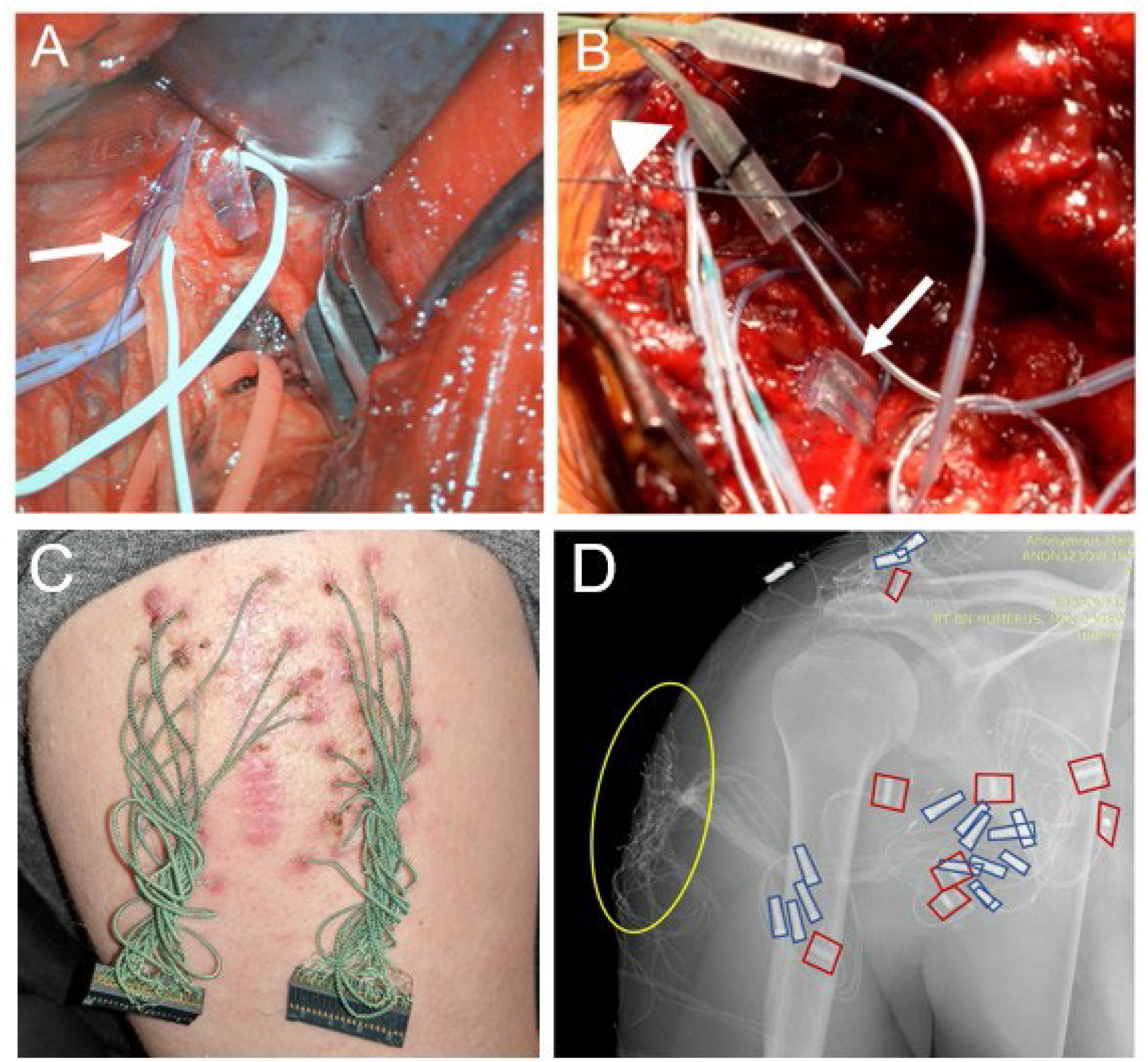
Surgical implantation of Composite Flat Interface Nerve Electrodes (C-FINEs) and tunneling of externalized leads. (A) C-FINE around the exposed radial nerve (not yet sutured closed, arrow). (B C-FINE on suprascapular nerve (arrow) and inline connector (arrowhead). (C) Inline connectors in nerve exposure incision (arrow) and tunneled externalized leads (arrowhead), which were subsequently individually tunneled to separate percutaneous exit sites. (D) X-ray of implanted nerve cuffs (outlined in red) and inline connectors (outlined in blue), with percutaneous exit sites (circled in yellow).

### Post-Op System Validation

Surgical outcomes were assessed through monthly visits to inspect wound healing and potential side effects or complications. We characterized the chronic recording (all) and stimulation (S1 only) capabilities of the implanted microelectrode arrays. We also measured the number of individually recordable neurons and their signal-to-noise ratios to quantify the efficacy of the recordings over a one-year period. To assess whether the recorded neural activity modulated with different attempted movements, the study participant was shown movements of different body segments and asked to attempt to perform those movements on cue^27,28^. Neural signals were recorded and analyzed to determine if, and at what point, the attempted movements were differentiable based upon neural activity alone. To assess the efficacy of the S1 arrays at producing sensation, microstimulation of individual contacts was delivered as 100 Hz trains of biphasic, cathode-first, charge-balanced pulses of 200 microsecond duration and 80 microamp amplitude (Cerestim R96, Blackrock Neurotech, Salt Lake City, UT). Stimulation of individual contacts of each C-FINE was used to determine the pattern of muscle activations achievable, as well as the muscle recruitment order. Muscle recruitment was determined by analyzing normalized electromyography (EMG) signals recorded through fine wire electrodes inserted under Ultrasound guidance into target muscles. Finally, we examined the joint movement responses to varying pulse amplitude and pulse width of several C-FINE contacts, as well as how simultaneous stimulation of multiple C-FINE contacts could result in formation of functional hand grasps.

## RESULTS

### Surgical Outcome

All surgical procedures were well tolerated by the participant with no significant procedural or long-term complications. There was no significant discomfort associated with the cranial or peripheral exit sites. The participant did not experience any new motor, sensory, or speech deficits.

### Motor Recording

Each channel on each microelectrode array could detect up to five individually-identifiable neurons, based upon their distinct wave shapes^29,30^ (Figure 4A). Higher neuron yields were seen on both S1, medial M1, and IFG arrays, compared with lower yields on the lateral M1 and AIP arrays. There were significant changes over time in the number of isolatable single neurons on each array, with decreases in neuronal counts seen on arrays implanted in somatosensory and primary motor areas and increases seen in arrays implanted in anterior intraparietal and inferior frontal gyrus areas (Figure 4B). In contrast, the average signal-to-noise ratio remained consistent over time (p>0.05, regression slope test) for all electrode arrays (Figure 4B). This data demonstrates the chronic health of the electrode-tissue interface remained stable up to one-year post-implantation.

**Figure 4.**
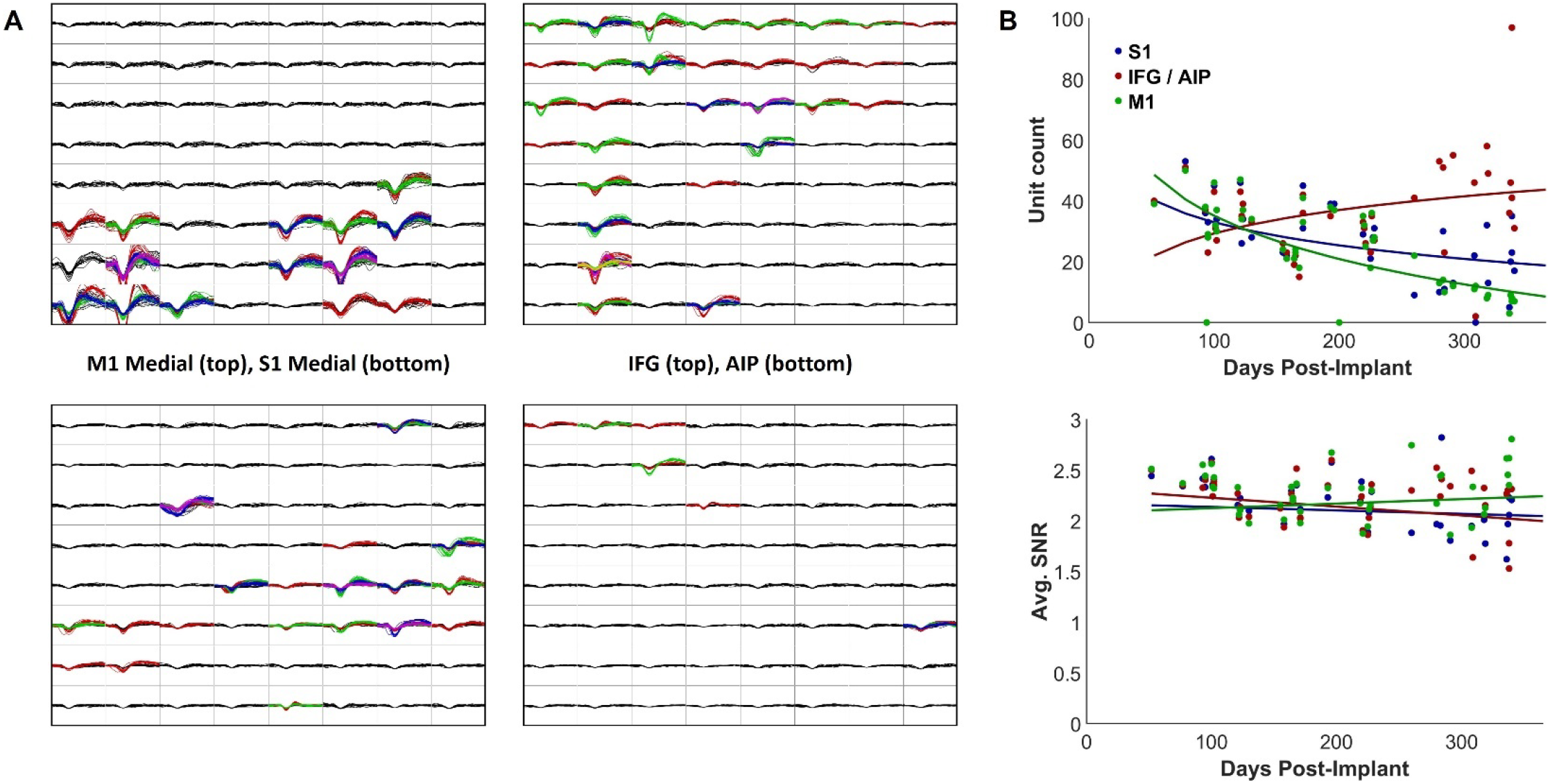
Cortical recording of individual neurons and stability over time. (A) Neural yields of the implanted arrays. Each 8×8 plot represents an implanted 64-channel microelectrode array, with each box insert showing the neural yield from a single channel. Channels could record multiple isolatable neural spikes (represented as different colors, data shown from post-implant day 123). Higher yields were observed on S1, M1, and IFG arrays, with lower yield observed on the AIP array. (B) Measurements of the chronic health of the electrode-tissue interface. Top plot: Number of isolatable neurons on each pedestal (n=128 channels per pedestal) over the first year of the study. Unit counts decreased over time on arrays implanted in S1 and M1, while they increased on the pedestal recording IFG and AIP (increases were mostly seen in IFG). The average signal-to-noise (SNR) ratio was relatively constant (p>0.05, regression slope test) across the one year of neural recordings (bottom plot).

We assessed whether the neural activity recorded from arrays in motor areas modulated to specific attempted movements. First, the participant was instructed to attempt to perform different hand grasps on cue, and the normalized multiunit spiking activity was recorded from arrays in motor-related areas. Each grasp pattern resulted in a different level of activation within individual electrodes (Figure 5A, sample electrode channel in IFG). This differential modulation to grasp pattern occurred about a second before the onset of the actual attempted movement and typically within a few hundred milliseconds after the cue. Second, the participant was asked to attempt to move eight different body parts. A classification algorithm correctly identified which body part was moved 79% of the time from recorded signals from the IFG array alone (Figure 5B). This diversity of body representations in what is conventionally thought of as ‘hand’ area is consistent with a previous study in the human hand knob area after paralysis^28^.

**Figure 5.**
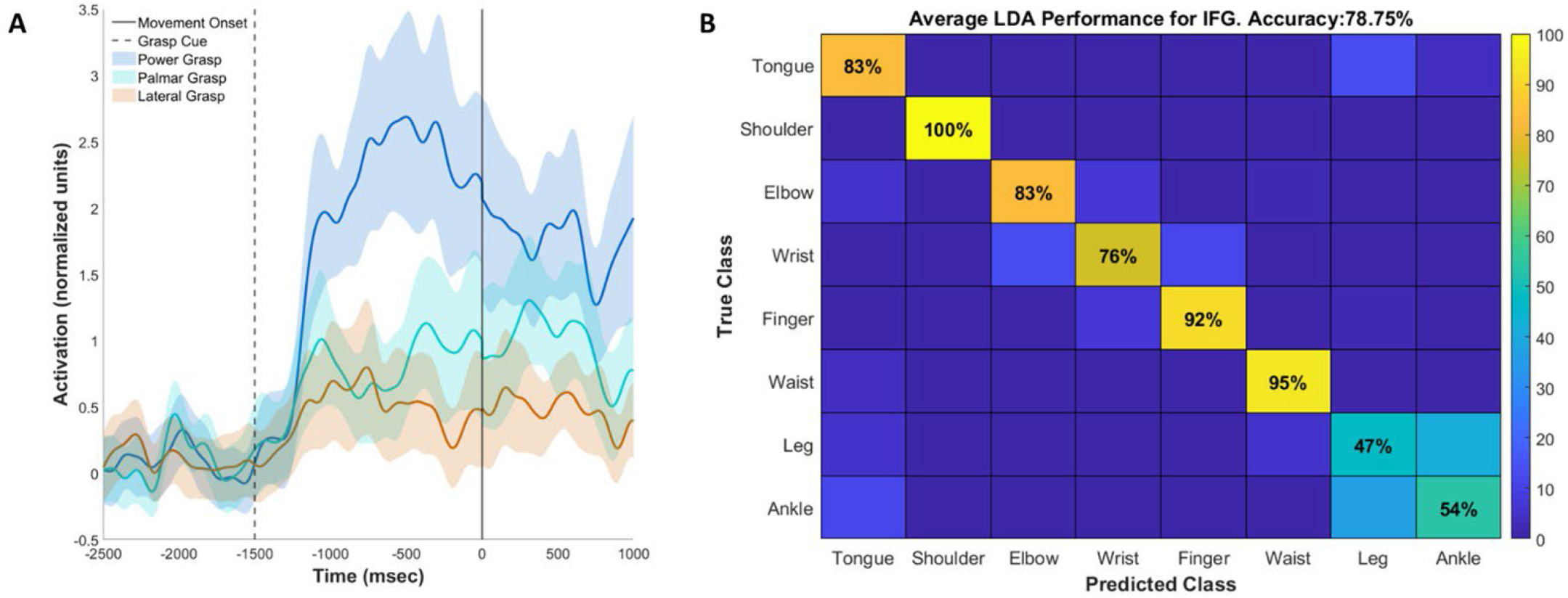
Modulation of neural signals to various attempted movements. (A) The participant attempted performing three different hand grasp patterns, which resulted in differentiable modulation in the neural activity. Shown is a sample electrode’s normalized mean activity timecourse (solid lines) and standard error (shaded regions) to imagination of power, palmar, and lateral grasp patterns. Imagination of grasp patterns resulted in distinguishable cortical responses both during the visually cued premovement phase (−1500 to 0 ms) and the imagined movement phase (after 0 ms). (B) Using cross-validated linear discriminant analysis, attempted movements of eight different body parts could be accurately classified in 79% of the trials from the neural activity on the IFG array alone.

### Sensory Stimulation

Stimulation through individual electrodes in the two 8×8 arrays implanted in the primary somatosensory cortex reliably produced sensation of touch on the index, middle, and ring fingers (Figure 6). Perceived locations typically included the anterior surface of the fingertips, and many projected fields also included the posterior side of the distal phalange(s). The microelectrodes eliciting reliable sensation tended to be along the posterior edge of the arrays. The locations of sensation elicited by intracortical microstimulation in each region of cortex (Figure 6) matched the sensory locations reported during intraoperative stimulation mapping of the same region (Figure 2A). Sensations were reported to feel similar to ‘pressure’ applied to the skin or ‘light squeezing’ of the fingertip.

**Figure 6.**
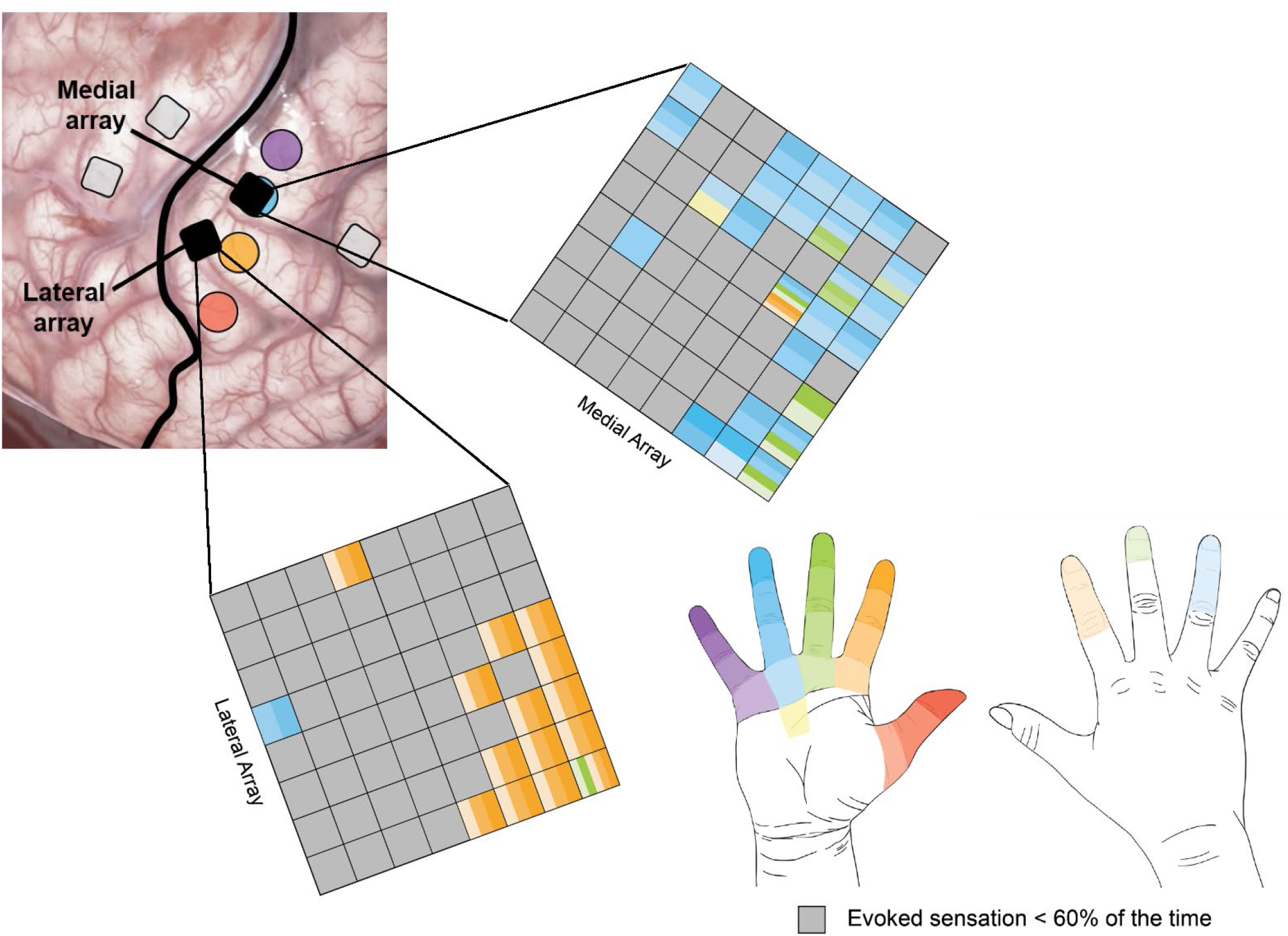
Sensory projected fields resulting from intracortical microstimulation applied to primary somatosensory cortex. Top left: the positions of the microelectrode arrays relative to the perceived locations reported during intraoperative mapping. Note that the locations reported were whole fingers rather than specific phalanges. Middle: The projected fields for each electrode in the medial (top) and lateral (bottom) arrays. The arrays are oriented to match their positions in the image of the cortex. Bottom right: Color legend for projected fields shown on the array diagrams. Each segment of the finger corresponds to a different shade. Grey electrodes may have elicited sensation in some trials, but the locations were not consistently reported for at least 60% of trials.

### Functional Electrical Stimulation

Stimulation of each C-FINE contact resulted in activation of muscles innervated by the stimulated nerve. As stimulation levels for a given C-FINE contact increased, additional muscles were recruited. While many contacts within a given C-FINE recruited similar groups of muscles, the order of recruited muscles differed across contacts (Figure 7). Recruitment order was more similar for neighboring C-FINE contacts, and recruitment patterns across contacts demonstrate the spatial clustering of muscle-specific efferents within the nerve fascicles (Figure 7). For example, stimulation of the radial C-FINE resulted in activation of wrist (ECRL) and finger extensors (EDC and EIP), thumb actuators (EPL and APL), and elbow flexion (BRR) and wrist supination (SUP). BRR activation was more prominent on contacts on the left half of the array while SUP activation was more prominent on the right half, and all other muscle activations were more distributed. Ulnar nerve stimulation produced activation of thumb flexion and adduction (FPB and ADP, predominantly on bottom half of the cuff), as well as index finger flexion (FDI, also predominantly on bottom half of cuff), with other functions (pinky abduction – ADM, finger flexion – FDP, and wrist flexion – FCU) distributed across the cuff contacts. The median nerve C-FINE produced wrist flexion (FCR – predominantly top half), forearm pronation (PT – predominantly right half), with finger (FDS, FDP) and thumb (FPL) flexion distributed across the contacts. Each of the musculocutaneous (biceps, brachialis), lateral pectoralis (upper and lower pectoralis) and axillary (anterior, middle, and posterior deltoid) nerve cuffs also produced spatially organized recruitment of their respective muscles.

**Figure 7.**
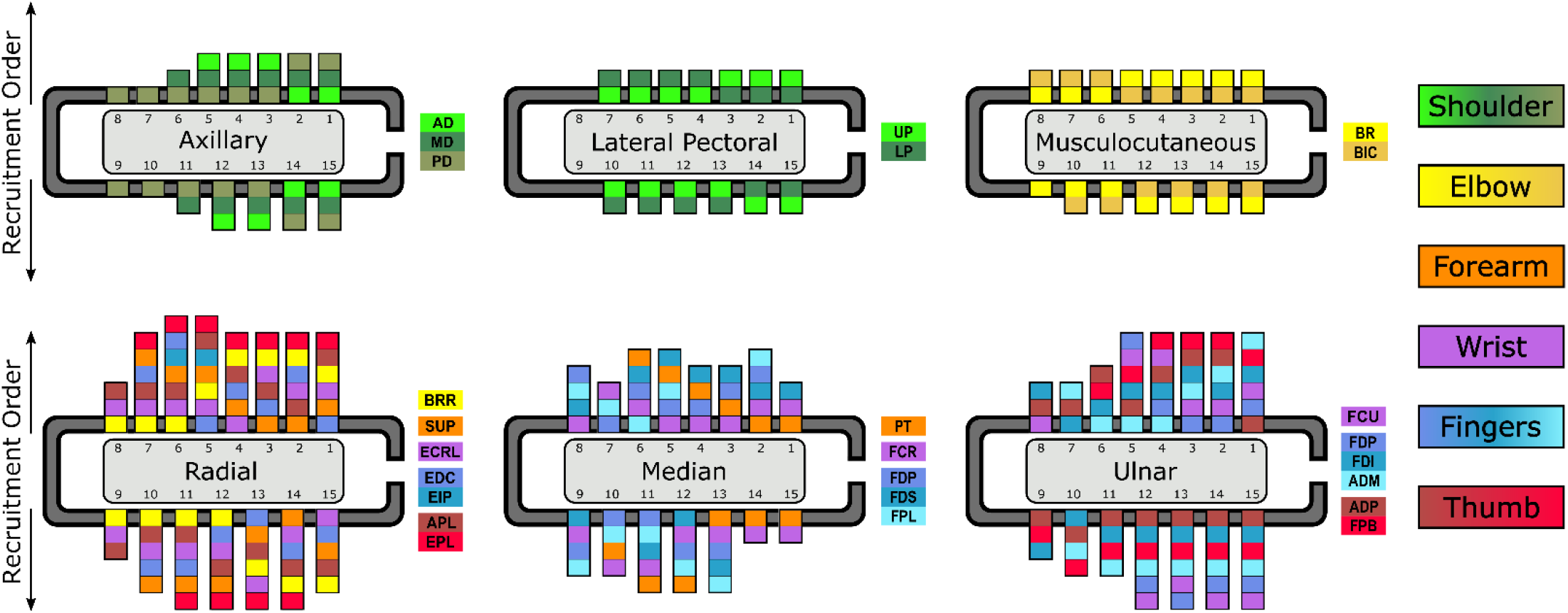
Muscle recruitment order associated with stimulation of individual contacts in the C-FINEs^37^. Muscles represented by a given color act on that color’s associated body part (e.g. all blue muscles activate finger flexion or extension) Several contacts exhibited spatial organization of muscle activation (e.g. left contacts of radial nerve more readily recruited brachioradialis, bottom contacts of ulnar nerve more readily recruited thumb muscles (ADP and FPB). Six of the eight implanted C-FINES resulted in activation of the expected set of muscles.

Nerve stimulation patterns involving multiple contacts on multiple nerves resulted in functional movements of reaching and grasping. Cortical signals related to the intended movement were decoded into a command signal, which then governed the level of stimulation delivered to sets of contacts in the C-FINEs (Figure 8). These patterns of nerve stimulation were able to generate the intended movement in the participant’s own limb. For example, a combined stimulation pattern of contacts on the median, radial, and ulnar nerves resulted in open/close of a lateral hand grasp, and the desired hand aperture was controlled by modulating the stimulation pulse width (Figure 8).

**Figure 8.**
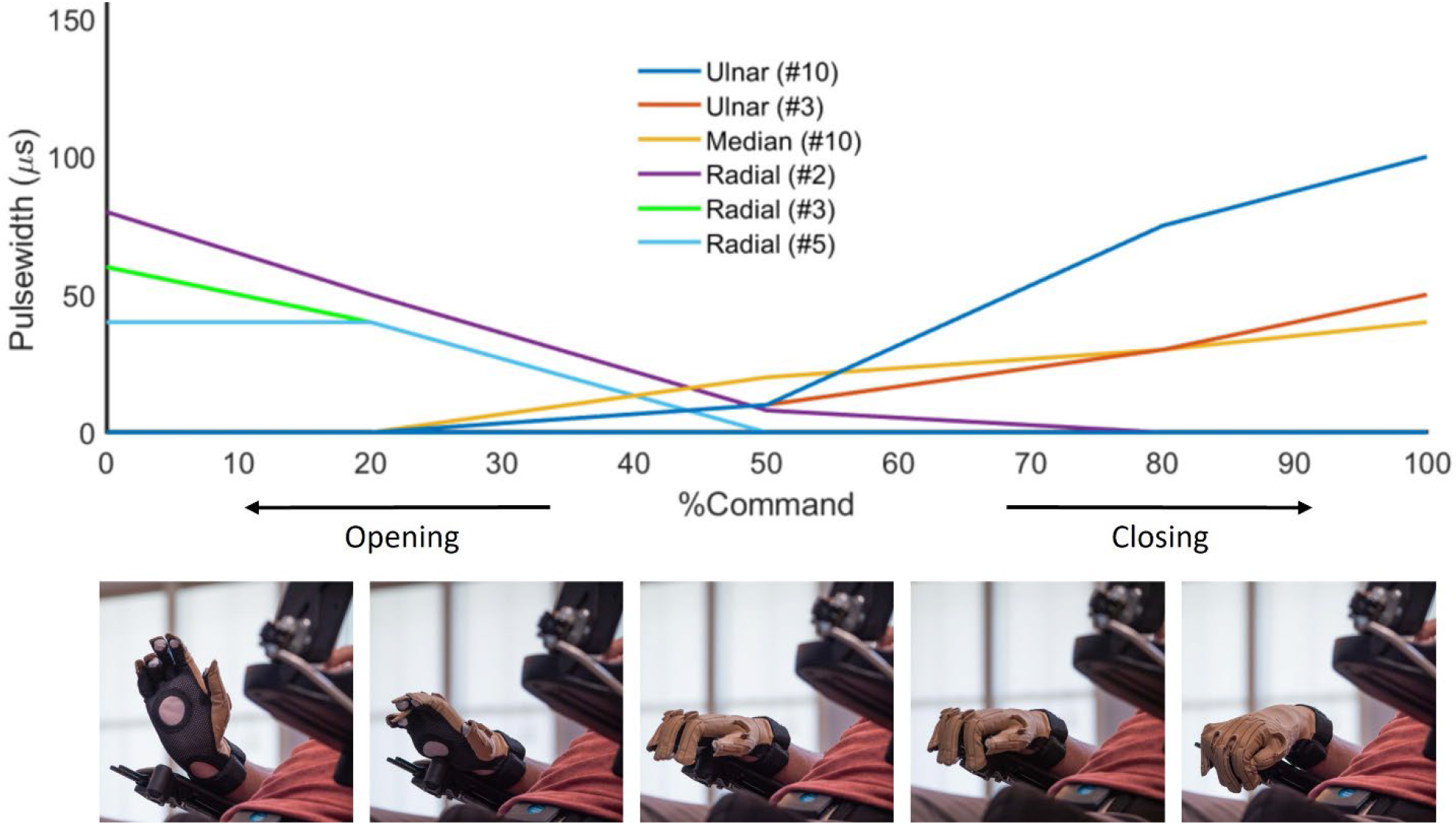
Simultaneous stimulation of sets of C-FINE contacts resulted in functional movements of the upper limb (hand shown here). Top: The desired hand state (x-axis) modulated the pulse width of electrical stimulation (y-axis) delivered to specific contacts on multiple nerves to produce the desired aperture. Bottom: Nerve stimulation resulted in varying degrees of opening (left) to closing (right) of the participant’s own hand.

## DISCUSSION

In this report, we describe a system for brain-controlled stimulation of peripheral nerves to restore voluntary hand and arm movement in a person with chronic motor-complete tetraplegia. We also demonstrate feasibility of a sensory cortical interface that provides touch percepts of the fingertips that can be used for sensory feedback during brain-controlled restored movement.

The most important advantage of the ReHAB system compared to other BMI systems, which have provided control of computer cursors or robotic limbs, is that it restores movement of the user’s own arm and hand using functional electrical stimulation (FES). FES has been used for many years to activate paralyzed muscles in people with neurological deficits^6,7^. In the lower extremity, FES systems have been used for standing, transfer, and walking applications^9,10^. In the upper extremity, FES systems have been shown to restore grasping functions to persons with cervical spinal cord injuries^6,8,31^. However, command options for telling the system what movements to make have been limited in the high tetraplegia population. The ReHAB system overcomes this challenge by using intracortical microelectrodes to provide a source of complex movement commands in this population.

An additional advancement in FES offered by the ReHAB system is the use of nerve cuff electrodes to activate motor efferents, rather than directly stimulating muscles with electrodes placed in individual muscles. Multi-contact cuffs can selectively activate different fascicles within a nerve^26,31,32^, and thereby selectively recruit one or more muscles innervated by a given nerve. Because a single cuff can recruit all muscles innervated by a given nerve, fewer interfaces need to be implanted to achieve full hand and arm function, thus reducing the necessary number of incisions and the complexity of the implantation procedure. In addition, since axons responsible for activation of synergistic muscles tend to be grouped into distinct nerves, peripheral nerve stimulation makes it possible to leverage the natural anatomic organization of the nerves to produce coordinated joint movements. Nerve stimulation can also achieve muscle activation at lower currents than direct muscle stimulation. By using multi-contact peripheral nerve stimulating electrodes rather than direct muscle stimulation, we were able to produce coordinated movement across a variety of muscles using relatively few electrode arrays and low stimulation intensities.

The ReHAB system expands on prior FES studies by providing pathways for direct recording of brain signals to control the patterns of peripheral nerve stimulation to restore voluntary movement. In addition to the potential for high density of information transfer, recording directly from motor areas of the brain may tap into native motor intentions, thus producing more intuitive control. In contrast to implantation only in primary motor areas of the brain, there are many potential advantages to recording from multiple upstream and downstream sensorimotor regions. In nonhuman primates, recording from ventral premotor regions have demonstrated neural activity that tunes to goal-directed activity rather than the more body-centric activity seen in primary cortex^33,34^. Likewise, in monkey and human experiments, single neurons from the anterior intraparietal area have been found to be selective for a variety of imagined hand postures that can be used to control prosthetic hands^23,35^. Combination of information from multiple distinct brain regions may allow for more intuitive and reliable motor control, and we have established that implantation in multiple brain regions is safe and well tolerated.

Finally, the ReHAB system combines brain-controlled arm movements with sensory feedback provided via microstimulation of primary somatosensory cortex. Sensory feedback is a critically important component of naturalistic movements of the upper extremity and could enhance object grasp and manipulation. Several prior studies have shown that focal brain stimulation can produce artificial sensory percepts and that these percepts can improve control of brain-controlled robotic limbs^18,36^, but prior systems have not combined this sensory feedback with brain-controlled FES. Notably, brain stimulation in our participant conveyed a naturalistic sense of touch on regions of the fingers critical for grasp function. Our results confirm that microelectrode stimulation can achieve sensory percepts at the level of individual fingertips, and demonstrate that sensory locations evoked by microelectrode stimulation are spatially congruous with those reported during intraoperative awake mapping.

### Limitations

There are several important limitations to this approach. First, only upper motor neuron injuries can be addressed using peripheral nerve stimulation, since denervation atrophy associated with peripheral nerve injury (including at the level of spinal cord injury) will preclude muscle activation. Second, it is necessary that the brain cortex be intact for recording of motor function, which prevents application when pathology involves damage to motor neurons (such as stroke) or progressive neurodegenerative conditions. Finally, this report involves a single participant, so additional studies will be necessary to determine whether the findings are generalizable.

## CONCLUSION

Brain-controlled peripheral nerve stimulation has the potential to allow restoration of arm and hand function after neurological injury. Use of a system like this to control the upper extremity may allow restoration of naturalistic movements, and future research may allow use of the reanimated upper extremity to perform simple activities of daily living.

## Data Availability

All de-identified data produced in the present study are available upon reasonable written request to the authors.

## Acknowledgments

We would like to thank the University Hospitals Cleveland Medical Center (UHCMC) Plastic Surgery Team for their expertise during the implant surgeries, and the UHCMC Clinical Core Staff for continued clinical support of the study participant. We thank Dr. David Preston for his expertise with intraoperative ultrasound. Finally, we thank the study participant (RP1) and family for their continued dedication to this work.

## Abbreviations

BMI: brain-machine interface
FES: functional electrical stimulation
SCI: spinal cord injury
ICMS: intracortical microstimulation
EMG: electromyography
AD: anterior deltoid
MD: medial deltoid
PD: posterior deltoid
UP: upper pectoral
LP: lower pectoral
BR: brachialis
BIC: biceps
BRR: brachioradialis
SUP: supinator
ECRL: extensor carpi radialis
EIP: extensor indicis proprius
EDC: extensor digitorum communis
APL: abductor pollicis longus
EPL: extensor pollicis longus
PT: pronator teres
FCR: flexor carpi radialis
FDP: flexor digitorum profundus
FDS: flexor digitorum superficialis
FPL: flexor pollicis longus
FCU: flexor carpi ulnaris
FDI: first dorsal interosseous
ADM: abductor digiti minimi
ADP: adductor pollicis
FPB: flexor pollicis brevis

